# Clinical Assessment of the Utility of Metagenomic Next-Generation Sequencing in Pediatric Patients of Hematology Department

**DOI:** 10.1101/2020.05.28.20115337

**Authors:** Heping Shen, Diying Shen, Hua Song, Xueqin Wu, Cong Xu, Guangyu Su, Chao Liu, Jingying Zhang

**Affiliations:** The Children’s Hospital, Zhejiang University School of Medicine, National Clinical Research Center for Child Health. Hangzhou, Zhejiang, China 310003; Hangzhou Matridx Biotechnology Co., Ltd, Bd 5, 208 Zhenzhong Rd, Hangzhou, Zhejiang, China 310030

**Keywords:** Metagenomic Next-Generation Sequencing, Diagnosis, Pediatric, Hematological malignancy, Infectious disease

## Abstract

Metagenomic Next-Generation Sequencing (mNGS) is an emerging technique for microbial identification and diagnosis of infectious diseases. The clinical utility of mNGS in management of infections, especially its real-world impact on antimicrobial therapy and patient outcome has not been systematically evaluated. To that end, we prospectively assessed the effectiveness of mNGS in 70 febrile inpatients with suspected infections at Hematology department of the Children’s Hospital, National Clinical Research Center for Child Health. 69/70 patients were given empirical antibiotics prior to mNGS. A total of 104 samples (62 plasma, 34 throat swabs, 4 bone marrow, 4 bronchoalveolar lavage) were collected on day 1–28 (mean 6.9) after symptom onset and underwent mNGS testing. Traditional microbial tests such as culture identified causal microorganisms in 5/70 (7.14%) patients that were in accordance with mNGS. In addition, mNGS reported possible pathogens when routine tests were negative. Antimicrobial treatment was adjusted in 55/70 (78.6%) patients that led to improvement/relief of symptoms within 3 days. On the other hand, in 15/70 (21.4%) patients, mNGS reports were considered irrelevant by a board of clinicians based on biochemical, serological, imaging evidence and experiences. We concluded that mNGS not only expanded clinical capacity of pathogen detection, but also yielded positive impact on handling suspected infections through 1) differential diagnosis which may rule out infectious diseases and 2) modification and de-escalation of empirical antibiotic regimen.

## Introduction

Clinical management of microbial infection is a leading factor that influences prognosis of pediatric patients with hematological malignancies including acute lymphoblastic leukemia (ALL), acute myeloid leukemia (AML), non-Hodgkin lymphoma (NHL), aplastic anemia (AA), blastic plasmacytoid dendritic cell neoplasm (BPDCN) [1–3]. Early diagnosis and intervention of infectious diseases would drastically reduce the risk of complications and improve patients’ long-term survival rate [4]. As a result, diagnostic tests play a critical role in clinical care of hematological patients with suspected infections, especially when empirical antibiotic treatment is not effective. However, traditional microbiological techniques such as culture, staining and serology often have low sensitivity and can only cover a small fraction of potential pathogens [5].

With the development of Massively Parallel Sequencing (MPS) technologies, sequencing-based diagnostics such as the metagenomic Next Generation Sequencing (mNGS) are increasingly applied clinically. mNGS test is “culture-free” and “hypothesis-free” that can encompass a wide range of pathogens in a single test [6]. Compared to contemporary microbiological methods, mNGS has the advantage of diagnosing polymicrobial infections, fastidious and unculturable or even novel pathogens within a time frame of 24–48 hours [7]. As a nucleic acid test (NAT), mNGS has fairly high sensitivity and specificity and is generally less-affected by antibiotic use than culture [8,9].

Despite its technological potential, mNGS has several drawbacks that might hinder its application in a clinical setting. These include but are not limited to high cost, slow turn-around-time (TAT), lack of assay standards and challenges in interpretation of reported microorganisms. Clinicians’ opinions towards the utility of mNGS are quite polarized. While many studies pointed out its value in diagnosing infections in immunocompromised cohort, a number of studies suggested limited utility of mNGS as currently used in routine clinical practice [10, 11]. The divergent views on mNGS and its real-world impact partly reflect the complexity of infectious diseases management and how to integrate mNGS into patient treatment. There are many factors at play, such as sample type, TAT, prior use of antibiotics, prevalence of antimicrobial resistance, patient’s age, ethnicity, gender, underlying illness, *etc*. [12]

In order to experience and assess whether and how mNGS can benefit patient outcomes, we incorporated mNGS as a routine microbiological test in the Department of Hematology at Children’s Hospital in Zhejiang Province, China. A total of 70 febrile pediatric patients with varying hematological diseases participated in this study. Most subjects were immunocompromised due to chemotherapy, hematopoietic stem cell transplantation and underlying conditions. Samples were collected and sent out for mNGS testing with most reports returned in less than 24 hours. For 38 patients who exhibited signs of respiratory infection (cough, shortness of breath, chest pain *etc*.), both respiratory sample (throat swab or bronchoalveolar lavage (BALF)) and peripheral blood were obtained. Routine clinical examinations such as chest CT scan, microbiological culture, blood cell count, biochemistry (PCT, hs-CRP, cytokines), serology (Flu A, Flu B, HPIV, AdV, RSV, EBV and CMV) and molecular (*Mycoplasma pneumoniae*) tests were performed in conjunction with mNGS. Prior to mNGS, 69/70 patients were empirically administered broad-spectrum antibiotics, but no apparent symptom relief was observed. The empirical antibiotic regimen typically comprised of meropenem, linezolid and voriconazole that targeted gram-negative, gram-positive bacteria and fungi, respectively. Antifungals were routinely used as a prophylaxis in hematological patients, which was deemed as a gold standard for preventing invasive fungal infections [13].

To interpret whether microorganisms reported by mNGS were of clinical relevance, we decided to evaluate the results according to the following criteria: we considered findings to be clinically relevant if 1) mNGS detected the same pathogens as shown in microbial culture/PCR; 2) patient’s symptoms improved within 3 days upon adjusting or de-escalating antibiotics based on mNGS; 3) mNGS discovered pathogens that have already been covered by existing antibiotics and patient’s symptoms relieved within 3 days; 4) mNGS was negative and final diagnosis indicated diseases not associated with microbial infection (*i.e*. drug fever, tumor, autoimmune diseases *etc*.). On the other hand, we considered mNGS results to be irrelevant if 1) it reported bacteria or fungi but the patient recovered by imposing antifungal or antibacterial treatment, respectively; 2) mNGS reported potential pathogens but no improvement was observed by altering antibiotics. In cases where more than one type of sample was tested (i.e. throat swab and blood), reports from both samples were compared and taken into consideration.

## Methods

### Subject recruitment

From May 2019 to October 2019, 70 patients (43 males, 27 females) at Hematology department of the Children’s Hospital, Zhejiang University School of Medicine, National Clinical Research Center for Child Health were recruited for this study. All participants were suspected of having infectious diseases due to fever and/or respiratory/gastrointestinal symptoms such as cough, shortness of breath, chest pain, diarrhea and abdominal pain *etc*. All subjects have provided informed consent for participation and publication of the de-identified data.

### Sample collection and mNGS testing

For each patient, 10 mL of peripheral blood was drawn into EDTA tube. Sample collection took place on day 1–28 (mean 6.9) following onset of fever. Throat swab or 10 mL of BALF was collected if respiratory symptoms were present. In 4 patients, bone marrow aspiration was performed, and 2 mL of bone marrow was acquired instead of blood. Following collection, all samples were immediately sent to Hangzhou Matridx Biotechnology Co., Ltd. for mNGS testing (PDC-seq^TM^). Whole blood was centrifuged at 1600g for 10 min and supernatant was centrifuged at 16000g for 10 min to obtain plasma. For plasma and bone marrow, DNA sequencing was performed. For throat swab and BALF, RNA sequencing was also performed in order to detect RNA viruses, which are common in community-acquired respiratory infections. Following nucleic acid extraction, DNA or RNA library was prepared by reverse transcription (RNA only), enzymatic fragmentation (except for plasma), end repairing, terminal adenylation and sequencing adaptor ligation (NGSmaster^TM^ library preparation, Matridx, Cat# MAR002). Sequencing libraries were quantified by real-time PCR (KAPA) and pooled. Shotgun sequencing was carried out on illumina Nextseq^TM^ platform. Approximately 20 million of 75bp single-end reads were generated for each library. Bioinformatic analysis was conducted as described in a previous report [14] in which sequences of human origin were filtered (GRCh38.p13) and the remaining reads were aligned to a microbial reference database (NCBI GenBank and in-house curated microbial genomic data) in order to identify species and relative abundance. For each sequencing run, one negative control (artificial plasma mixed with fragmented human genomic DNA) and one positive control (a mixture of inactivated bacteria, fungi and pseudoviral particles containing synthesized DNA or RNA fragments of adenovirus and influenza A virus, respectively) were included.

## Results

### Clinical characteristics

In this study, 51/70 (72.9%) participants were under 10 years old. All patients have been previously diagnosed with hematological diseases. In particular, 46 patients had acute lymphoblastic leukemia (ALL), 8 had acute myeloid leukemia (AML), 13 had non-Hodgkin lymphoma (NHL), 1 had aplastic anemia (AA), 1 had blastic plasmacytoid dendritic cell neoplasm (BPDCN) and 1 had immunodeficiency disease (IDD). Patients’ samples were collected for mNGS testing at day 1–28 (6.9 ± 4.9) following symptom onset (**Table 1** and **Supplemental Table 1**). Results of clinical examinations were shown in **Table 2**. Notably, 36/70 (51.43%) had abnormally low neutrophil count (neutropenia), which is typical in hematological patients [15].

**Table 1.** Clinical characteristics of study participants.

|  |  | Count | Ratio |
| --- | --- | --- | --- |
| Gender | Male | 43 | 61.40% |
|  | Female | 27 | 38.60% |
| Age (years) | $\geq 10$ | 19 | 27.10% |
| | $< 10$ | 51 | 72.90% |
| Sample type | Blood | 62 | 59.62% |
|  | Nasopharyngeal swab | 34 | 32.69% |
|  | Bronchoalveolar lavage | 4 | 3.85% |
|  | Bone marrow | 4 | 3.85% |
| Underlying disease | ALL | 46 | 65.71% |
|  | NHL | 13 | 18.57% |
|  | AML | 8 | 11.43% |
|  | AA | 1 | 1.43% |
|  | BPDCN | 1 | 1.43% |
|  | IDD | 1 | 1.43% |
| Neutropenia | Agranulocytosis | 36 | 51.43% |
|  | Normal | 34 | 48.57% |
| Chest CT scan | Abnormal | 30 | 42.86% |
|  | No abnormalities | 40 | 57.14% |
| Clinical outcome | Recovered | 69 | 98.57% |
|  | Deceased | 1 | 1.43% |

Abbreviations: acute lymphoblastic leukemia (ALL), acute myeloid leukemia (AML), non-Hodgkin lymphoma (NHL), aplastic anemia (AA), blastic plasmacytoid dendritic cell neoplasm (BPDCN), immunodeficiency disease (IDD).

**Table 2.** Routine laboratory test results.

| Test | Count |
| --- | --- |
| WBC | 0.01 - 19.14 (3.38) x10<sup>9</sup>/L |
| Neutrophill | 0 - 10.9 (1.63) x10<sup>9</sup>/L |
| hs-CRP | 0.5 - 177.88 (43.42) mg/L |
| PCT | 0.065 - 2.43 (0.36) ng/mL |
| IL-6 | 4 - 397.1 (73.17) pg/mL |
| IL-10 | 2.2 - 266.2 (18.74) pg/mL |
| γ-INF | 1 - 116.3 (10.28) pg/mL |
| TNF | 1 - 24.6 (2.53) pg/mL |

### Microbiological testing and clinical impact

Blood culture and *Mycoplasma pneumoniae* (MP) RNA test reported positive findings in 5/70 (7.14%) patients. Particularly, blood culture identified 1 case of *Pseudomonas aeruginosa*, 1 case of *Klebsiella pneumoniae* and 1 case of *Staphylococcus epidermidis* while MP RNA test identified 2 positive cases (1 from throat swab and the other from BALF). These microbial findings were also reported by mNGS (**Supplemental Table 1**). Serology tests identified 3 case of EBV (1/3 reported by mNGS), 2 case of CMV (1/2 reported by mNGS), 1 case of *Aspergillus fumigatus* (not reported by mNGS), 1 case of HPIV (not reported by mNGS since RNA sequencing was not performed for plasma).

In total, 104 samples (62 plasma, 34 throat swabs, 4 bone marrow, 4 BALF) were tested. mNGS on 47/62 plasma, 34/34 swab, 3/4 bone marrow, 4/4 BALF reported positive microbial findings, respectively (**Figure 1**). For 34 patients whose blood and swab were both tested, mNGS detected overlapping microorganisms in 14/34 (41.18%) patients, including human herpesviruses (5 cases of EBV, 1 case of HSV-1, 1 case of HHV-7), human parvovirus B19 (3 cases), *Pseudomonas aeruginosa* (2 cases), *Acinetobacter baumannii* (1 case), *Staphylococcus aureus* (1 case), *Haemophilus parainfluenzae* (1 case) and *Candida albicans* (1 case). For 4 patients whose blood and BALF were collected, mNGS reported overlapping microorganisms in 2/4 (50%) (1 case of *Mycoplasma pneumoniae* and 1 case of human parvovirus B19) (**Supplemental Table 1**).

**Figure 1.**
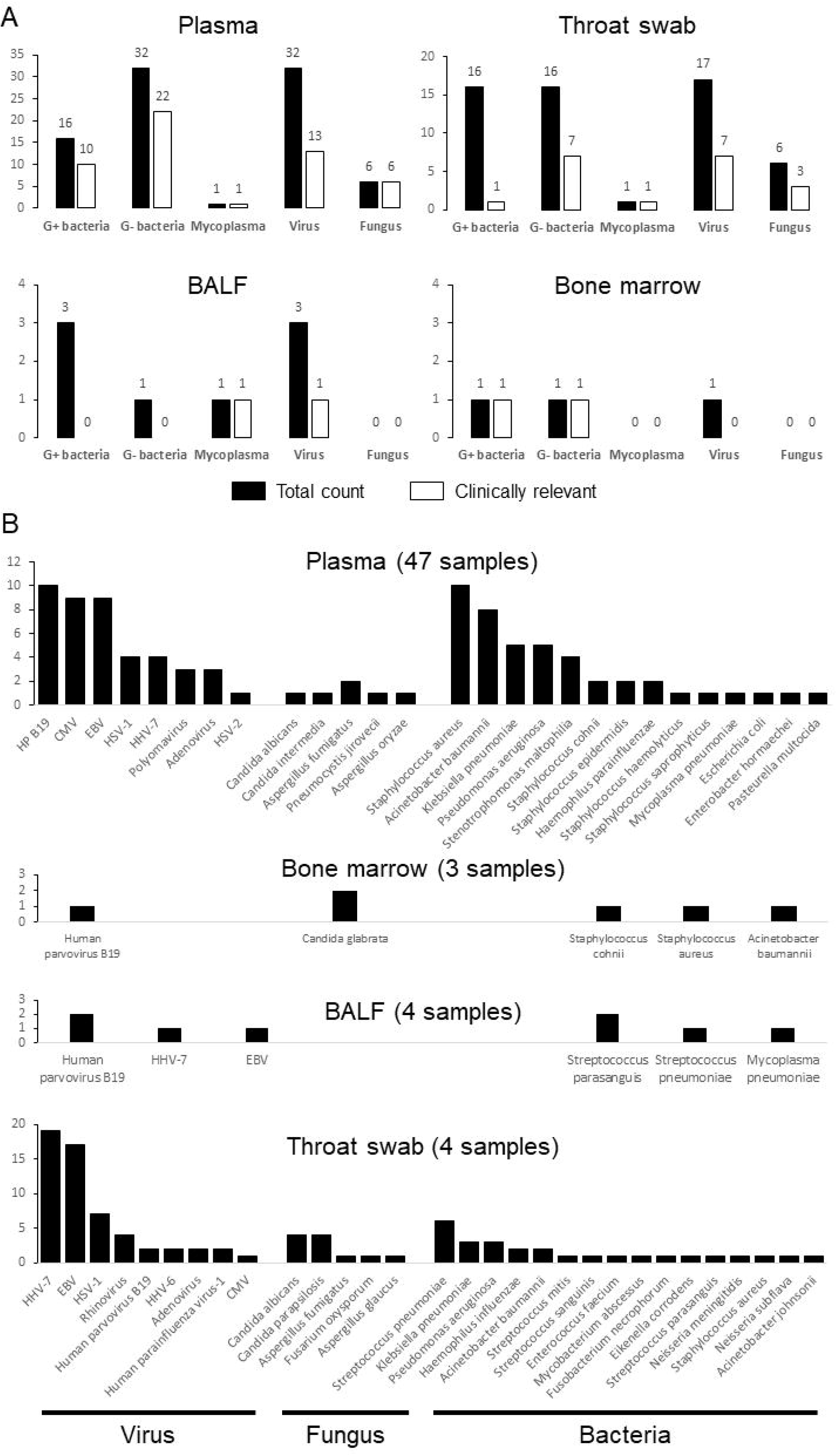
mNGS reports and clinical relevance. A total of 104 samples (62 plasma, 34 throat swabs, 4 bone marrow, 4 BALF) were collected for mNGS testing. 47/62 plasma, 34/34 swab, 3/4 bone marrow, 4/4 BALF reported positive microbial findings, respectively. The total counts of microorganisms of different categories and the number of microorganisms deemed clinically-relevant were shown (A) while the species information and count of viruses, fungi and bacteria detected from 4 different sample types were graphed (B).

In 55/70 (78.57%) patients, mNGS findings led to positive impact on patient outcome through adjusting or de-escalating empirical antibiotics. Notably, for two patients with osteomyelitis, all microbial tests were negative except that plasma mNGS reported *Staphylococcus aureus*. Both patients recovered after applying anti-G+ bacteria treatment, including Vancomycin and Linezolid. In another patient with recurrent fever, plasma mNGS reported *Staphylococcus aureus* when routine tests were inconclusive. We adjusted antibiotics accordingly and the patient’s fever subsided. However, fever recurred after drug withdrawal and a second blood culture was able to identify *Staphylococcus aureus*, which was consistent with the earlier mNGS result. Moreover, in two cases where mNGS was negative, one patient was diagnosed with hemophagocytic histiocytosis and the other was diagnosed with myelodysplastic syndrome, both of which were not associated with microbial infection. Antibiotics were de-escalated based on mNGS results in both cases.

On the other hand, in 15/70 (21.43%) patients, mNGS either reported microorganisms that were deemed irrelevant (1 case of neoplastic fever, 2 cases of graft rejection after hematopoietic stem cell transplantation, 3 cases of bacterial enteritis) or failed to identify the causal pathogens when other tests and clinical evidence strongly suggested microbial infection (5 cases of pneumocystis pneumonia, 2 cases of pulmonary aspergillosis) (**Supplemental Table 1**).

## Discussion

### Clinical value of mNGS in diagnosing infectious disease

Failure or delay in identifying the microorganisms responsible for critical infection could lead to prolonged hospitalization and elevated death rate [16]. Patients with hematological disorders are immunodeficient due to chemotherapy, hematopoietic cell transplantation and therefore are prone to opportunistic infections. In these patients, the causal pathogens may consist of a wide range of microbes, including bacteria, fungi and viruses. Due to frequent use of broad-spectrum antibiotics and clinical prevalence of fastidious microorganisms, microbial culture often fails to detect the culprit. Typically, the positive detection rate of blood culture is less than 10% [17]. In this study, the detection percentage was 5.71% (4/70). In contrast, mNGS is culture-independent and hypothesis-free that can theoretically detect over 10000 pathogens of known genomic sequences. The technical detection rate of mNGS (percentage of mNGS reports that show potential pathogens) in our study was 88/104 (84.62%), making it a valuable tool for diagnosis of polymicrobial and rare infections, especially for immunocompromised patients.

In addition to bacteria and fungi, mNGS identified a variety of DNA and RNA viruses from all sample types, including adenovirus, rhinovirus, parainfluenza virus, human parvovirus B19, polyomaviruses (Trichodysplasia spinulosa polyomavirus, JC virus, BK virus) and human herpesviruses (HSV-1, HSV-2, EBV, CMV, HHV-6 and HHV-7), which were of clinical value since we normally lacked diagnostic tests for these viruses (**Figure 1**). Prior to mNGS, antiviral treatment was not as much considered as compared to bacterial and fungal infections. Using mNGS, we were able to consider the possibility of viral infections and decide early on whether antivirals were necessary. In addition, the high incidence of herpesviruses as shown in our cohort was consistent with previous findings in which these viruses are common even in healthy individuals. However, it has been proposed that there exists a dynamic interaction between host immunity and herpesviruses that may influence the outcome of concurrent infections, cancers and grafts [18]. The clinical implications of herpesviruses in immunodeficient patients need further explorations.

### Sampling is critical for interpretation and utility of mNGS

Plasma mNGS was of greater significance in helping clinicians determine the causal pathogen(s) as compared with throat swab, in which commensal microbiota from upper respiratory tract were present. Cell-free nucleic acid in plasma is thought to originate from apoptotic human cells and shedding of microbial nucleic acid from site of infection through interactions between the host immune system and invading pathogens [19]. In our study, 38/70 (54.29%) patients were suspected of respiratory infection due to symptoms such as cough and shortness of breath. Indeed, previous studies also suggest that respiratory tract infection (RTI) is one of the most frequently-encountered type of infection in hematological patients [20]. While BALF was of great value in diagnosing pulmonary infections, performing bronchoscopy on pediatric patients is not as frequent as in adults. Hematological patients often have low platelet count, further limiting the application of invasive procedures. Moreover, plasma mNGS has a higher likelihood of yielding false-negative results due to interference of host DNA background. Taken together, when bronchoscopy was not an option, collecting both throat swab and blood appeared to be a working strategy. As shown in our study, matching organism(s) from throat swab and plasma were identified in 14/34 (41.18%) patients (highlighted in red, **Supplemental Table 1**). It would also benefit patients when blood mNGS showed negative results, since throat swab can serve as a “backup” under this circumstance.

### When to use mNGS

The turnaround time (TAT), cost and interpretation of results are important factors to consider before application of mNGS in a clinical setting. Furthermore, nucleic acid extraction seemed to be another key element to ensure sensitivity towards pathogens with thick cell walls or capsules that prevent efficient release of nucleic acid for downstream sequencing. This is especially evident when it comes to fungi as mNGS appeared to have missed 5 cases of pneumocystis pneumonia and 2 cases of pulmonary aspergillosis. Fungal cell walls comprise of N-acetylglucosamine polymer chitin and extraction techniques such as bead-beating and sonication might be desirable for effective detection of these pathogens.

To summarize, we found mNGS to be of clinical value and can benefit hematological patients in the following scenarios: 1) patients in critical condition with suspected infection and effective treatment is urgently needed; 2) patients with immunosuppression or neutropenia that make them more disposed to polymicrobial infections; 3) routine microbiological tests are repeatedly inconclusive and empirical antibiotic treatment is unsuccessful; 4) recurrent of persistent fever of unknown origin in which differential diagnosis is needed to confirm or rule out microbial infection.

## Data Availability

All data generated or analyzed during this study are included in this article.

---

**Supplemental Table 1. Clinical information of study participants.** Detailed clinical characteristics, laboratory test results, mNGS results and record of antibiotic use was listed for 70 patients. To protect the privacy of each participant, all identifying information was omitted.

